# Development of machine learning-based algorithms for classifying physical activity intensity using wrist and thigh worn wearables

**DOI:** 10.1101/2024.08.07.24311585

**Authors:** Nicholas A. Koemel, Raaj K. Biswas, Matthew N. Ahmadi, Emmanuel Stamatakis

**Author notes:** Equally contributing 1^st^ author (Nicholas A. Koemel, Raaj K. Biswas).

## Abstract

The use of semi-supervised learning approaches can be used to extend a base-level classifier and offers a significant advantage by reducing the need for extensive labeled datasets. We utilized a two-stage semi-supervised learning model to classify physical activity intensity for wrist and thigh worn monitors, by retraining a base classifier with free-living wearable sensor data. Data was collected in two-phases comprising a laboratory and free-living session. Total classified time spent in light intensity, moderate intensity, and vigorous intensity were not significantly different from ground-truth minutes for either placement. The machine learning classifiers re-trained on free-living data accurately predicted light, moderate and vigorous intensity between both device placements. These findings demonstrate that similar estimates of physical activity intensity can be correctly classified for wrist and thigh placements when using semi-supervised techniques.

## INTRODUCTION

Accurate and accessible information on daily physical behavior is crucial for advancing public health initiatives and guiding policy decisions concerning physical activity. To achieve this, precise measurement and classification of physical activity intensities are essential. Supervised machine learning approaches have provided alternative methods to classifying physical activity intensity from wearable sensor data. While supervised machine learning methods have provided valuable insights, model development and training requires datasets collected under diverse free-living conditions with labelled or ground-truth annotation. However, there is are considerable wearable sensor datasets collected under free-living conditions that are available but do not have ground-truth annotations. Thus, there is a pressing need to explore more advanced data processing techniques that can leverage the availability of both labelled and unlabelled datasets to develop generalizable machine learning intensity classification models to assess habitual physical activity levels.

In contrast to supervised machine learning, semi-supervised models offer a promising alternative for improving the classification of physical activity intensity. Pattern recognition techniques from supervised models such as decision trees, random forests, and support vector machines, have demonstrated their potential in enhancing the accuracy of activity type prediction and intensity assessment. Random forests, in particular, utilize an ensemble of decision trees to deliver robust classification results with minimal data pre-processing and effective feature selection. The use of semi-supervised learning approaches can be used to extend a base-level classifier (eg: Random Forest) and offers a significant advantage by reducing the need for extensive labeled datasets and accommodating diverse data sources.

This paper aims to address the limitations of current physical activity measurement methods by utilizing a two-stage semi-supervised learning model to classify physical activity intensity for wrist and thigh worn monitors. By retraining a base classifier with free-living wearable sensor data, this study seeks to enhance the accuracy of intensity classification. This methodology addresses the need for more robust activity intensity recognition in diverse free-living settings, thereby contributing to a more reliable and generalizable physical activity assessment.

## METHODS

### Study Design

This project contained a two-part data collection phase including 1) a single approximately 1.5-hour long laboratory data collection session and 2) 6-hour long session of free-living observation. Data from the laboratory sessions will be used to train a base intensity classifier using wrist and thigh-worn devices. The free-living observation will use a combination of unlabeled and labelled video data to retrain and validate our intensity classifier via a semi-supervised machine-learning approach.

### Participants

The core analysis included 46 adults (Mean age: 57.43 ± 12.30 years; 67.4% female). Participants were recruited via social media, flyers, and word of mouth. Inclusion criteria included being aged 30-75 and comfortable completing activities of normal daily living (e.g., walking, cleaning etc) without discomfort. Individuals were excluded from the study if they reported having a heart condition or major operation, consuming cardiovascular medication that interrupts normal cardiovascular function or heart rate (e.g., Beta-blockers, antiarrhythmics) or a recent muscle ligament or joint injury worsened by physical activity. All participants completed written consent forms before the sessions and Ethics approval was obtained from the Sydney Local Health District, Royal Prince Alfred Hospital.

### Laboratory Sessions

Standard anthropometric values including height and weight were collected from each participant before beginning the study. Participants then completed a 90-minute structured ‘daily living’ session which included completing 11 activities each lasting 5 minutes in duration. Each participant completed the activities wearing a portable indirect calorimetry unit (Cosmed K5; Rome, Italy) and a Polar heart rate monitor (Kempele, Finland) to capture intensity and energy expenditure. The activities included 1) Lying supine in a relaxed state; 2) Riding on public transport in a seated position; 3) Standing dish washing by hand; 4) Picking up household items from ground level and placing them on a table; 5) Vacuum cleaning; 6) Standing laundry folding; 7) Casual walking at a comfortable pace self-defined by the participant; 8) Brisk walk or jog self-defined by the participant; 9) Stair climbing; 10) Cycle; and 11) Machine and free weight exercises. The walking activity was completed as an unstructured outdoor walk or indoors on a treadmill. For the brisk walk or jog activity, participants were instructed to find a vigorous pace they are comfortable sustaining for the 3 to 5-minute period to meet vigorous intensity as indicated by the heart rate (>77-95% heart rate maximum) and indirect calorimetry measured V0_2_ (>6 METs). Participants completed each task under the supervision of two trained professionals. Following each activity, the participant was provided with a rest period to allow the heart rate to return to the observed resting rate. Further detail regarding each of the activities is provided in Supplementary Table 1.

### Instrumentation

This study included wearable device measurements from the Axivity AX6 (Newcastle, United Kingdom) placed on the dominant wrist secured via a wrist strap and a rubber encasing (Supplementary Figure 1). The thigh device was placed 10 cm above the proximal part of the patella (Supplementary Figure 2) and secured by a hypoallergenic transparent dressing (Tegaderm™, St. Paul, Minnesota). The Axivity AX6 collects acceleration measurements using a 6-axis motion sensor. This device captures angle and linear velocity using an integrated accelerometer and gyroscope and offers a sampling range of 12.5-1600Hz with a configurable dynamic range of ± 2-16 gravitational units (g). We initilised all devices in the present study to collect data at a sampling rate of 100Hz. Time and frequency domain features from the raw accelerometry and gyroscope data using non-overlapping 10 second windows. Supplemental Table 2 provides the full list of features that were extracted from the accelerometer and gyroscope.

### Free-Living Observation and Coding Procedure

A second sample of 48 participants (Mean age: 59.3 ± 12.1 years; 56.3% female) completed a Free-Living Environment data collection session to re-train and test our activity classification algorithms while allowing for natural activities and organic behaviour transitions^3^. A total of 48 participants wore the devices and a neck-worn video camera to capture ground-truth evidence of physical activity behaviours. For this sample, the subject wore a portable camera (Losoform Z02; Guangzhou, China) positioned on the chest using a body-mounted camera holder (Telesin; Shenzhen, China). Once the camera was mounted in the camera holder, the subjects were instructed to wear the device for the following 6 hours to capture normal day-to-day habits.

For half of the collected free-living sample (n = 24), video files collected from the subjects were imported into the Noldus Observer XT 16 software for continuous observational coding (Noldus Information Technology; Wageningen, The Netherlands). This sample was classified as the ‘labelled’ sample, and the remaining uncoded sample was classified as the ‘unlabeled sample. For the labelled sample, we created a custom observation scheme that included base-level physical activity compendium categories 1) Lie 2) Reclining 3) Sit 4) Walk 5) Run 6) Stair Climbing 7) Eating. If a participant was not in the view of the camera the behaviour was coded as “Out of view”^4^. Once imported into Noldus, the videos were coded continuously by an individual coder which generates a continuous time-series vector of behaviours. These time series were then used to establish overlap with the time-stamped accelerometry data and corresponding predicted activity classification. Before each study participant, a master time-piece was synchronized to the computer that initialized the devices to ensure the time series of the accelerometry corresponded with the start time of the free-living video files^4,5^. For the physical activity classifier, each of these activities was then classified as either reclining, sitting, standing, walking activities, running or high energetic activities, stair climbing, cycling, or weightlifting (Supplementary Table 1). For each of these activities, a predicted intensity was provided including sedentary behaviour, light activity, moderate activity, and vigorous activity. The interobserver and intra-observer reliability was examined by coding videos with one primary coder and two alternative coders, selected at random. The intraclass coefficient or the coding class activity for interobserver (n = 2: 12 hours of video data) was 0.84 (95% Confidence Interval (CI): 0.77-0.90). The intraobserver was measured with the primary observer (n = 1: 6 hours of video data) and had an intraclass coefficient of 0.97 (95%CI: 0.96-0.99).

For activity classification, we applied two separate approaches depending on the placement of the accelerometer. For thigh placement, we applied a previously validated MatLab-based software application known as Acti4^6,7^ which employs decision trees to classify each of the above activities. For wrist placement, we used a random forest model which operates by establishing a single node that splits into branches followed by additional nodes, all met with a new binary decision^9^. These decisions are informed by learning on a bootstrapped sample of training data, which in this case is a combination of laboratory and free-living data. The nodes and branches will continue to split until a stop condition has been met which is in this study was determined by the information gained index which offers information into the greatest reduction in entropy or label impurity^5,10^.

Following activity classification, this study follows a three-stage development plan to build and refine our intensity classifier. First, we trained our intensity classifier using the in-laboratory activities of daily living. This step includes using the extracted features from the raw accelerometry and gyroscope data in combination with activity type, to classify movements as sedentary behaviour (SB), light intensity physical activity (LPA), moderate intensity physical activity (MPA), or vigorous intensity physical activity (VPA) in 10-second windows. Second, we applied a semi-supervised approach to re-train our base classifier using the coded or ‘labelled’ free-living data^11^. Third, we then tested this classifier using a sub-sample of unlabeled free-living data (n = 24). This included testing the classifier on the 6 hours of data available for each individual.

### Statistical Analysis and Classifier Performance

The classifier performance was evaluated using overall F1-scores and weighted kappa (*k*) coefficients. To explore the level of our classifier across the intensity of each activity, we also provide confusion matrices that include SB, LPA, MPA, and VPA. Bland-Altman plots used to examine mean bias and 95% limits of agreement (LOA) for time spent in each intensity category. Mixed effects linear regression with nesting of sex were used to compare intensity classification time.

## RESULTS

Participant characteristics are reported in Tables 1 and 2. Average age of the free-living sample was 58.8 (12.4) years and average age of the laboratory sample was 57.0 (12.5) years. Overall F1-scores, weighted kappa coefficients, and class-level sensitivity, specificity, and precision and heatmap confusion matrices are reported in. Across sex and age categories, the wrist classifier showed very good to excellent classification of sedentary behaviour (>84% across sex and age categories), moderate intensity (>86%), and vigorous intensity (>94%), and moderate to very good classification of light intensity (76% to 83%). The thigh classifier showed excellent classification of sedentary behaviour (>97%), very good to excellent classification of moderate intensity (>84%) and vigorous intensity (>92%), and moderate to very good classification of light intensity (72% to 86%). Total classified time spent in light intensity, moderate intensity, and vigorous intensity were not significantly different from ground-truth minutes for either placement. The bland-altman plots are shown in Figure 2. The difference between classified and ground truth minutes in sedentary, light, moderate, and vigorous indicate good agreement and no bias with increasing time for the wrist and thigh classifiers.

**Table 1:**
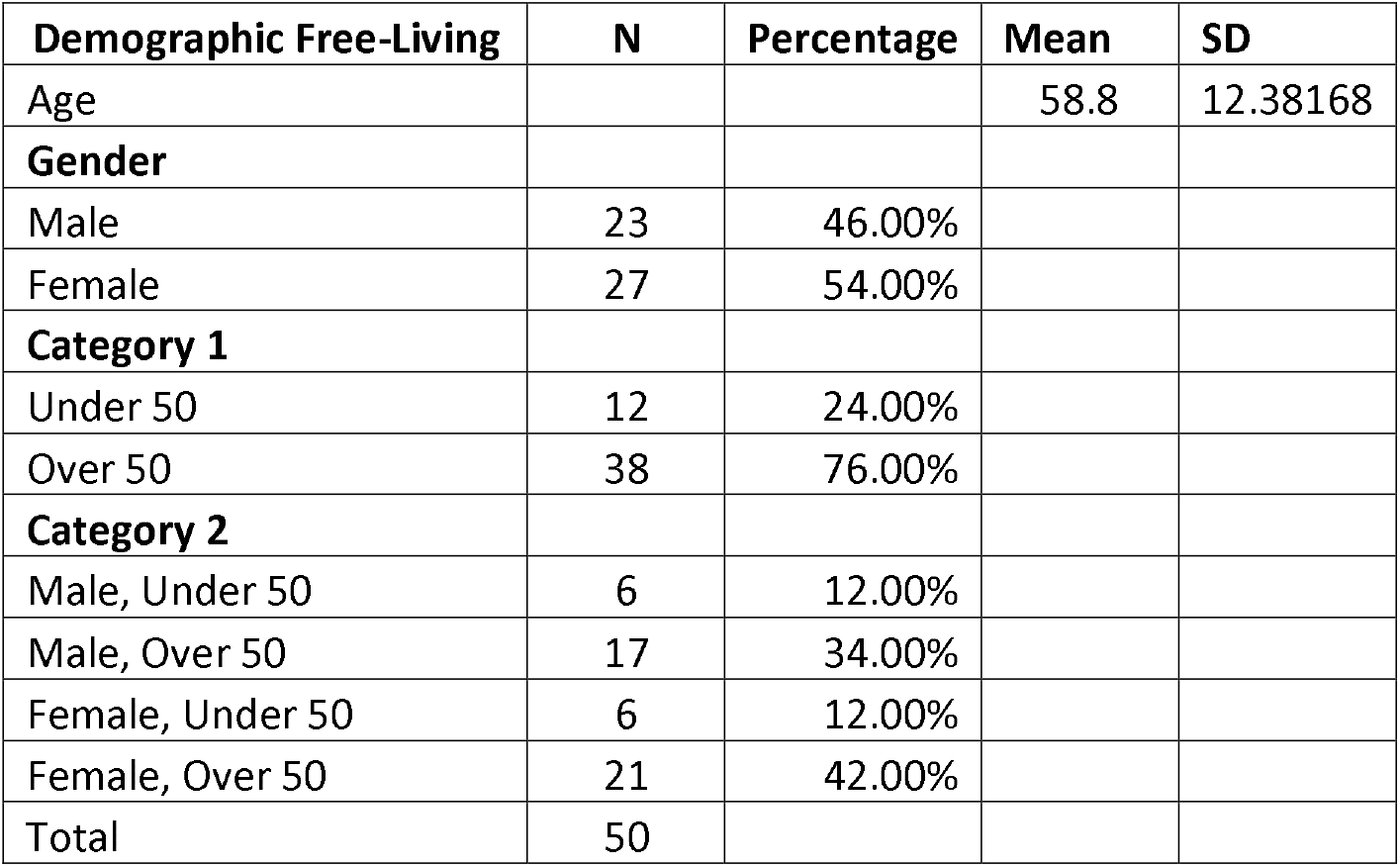

| **Demographic Free-Living** | **N** | **Percentage** | **Mean** | **SD** |
| --- | --- | --- | --- | --- |
| Age |  |  | 58.8 | 12.38168 |
| **Gender** |  |  |  |  |
| Male | 23 | 46.00% |  |  |
| Female | 27 | 54.00% |  |  |
| **Category 1** |  |  |  |  |
| Under 50 | 12 | 24.00% |  |  |
| Over 50 | 38 | 76.00% |  |  |
| **Category 2** |  |  |  |  |
| Male, Under 50 | 6 | 12.00% |  |  |
| Male, Over 50 | 17 | 34.00% |  |  |
| Female, Under 50 | 6 | 12.00% |  |  |
| Female, Over 50 | 21 | 42.00% |  |  |
| Total | 50 |  |  |  |

**Table 2.**

| **Demographic In-Lab** | **N** | **Percentage** | **Mean** | **SD** |
| --- | --- | --- | --- | --- |
| Age |  |  | 57.02083 | 12.51465 |
| **Gender** |  |  |  |  |
| Male | 17 | 35.42% |  |  |
| Female | 31 | 64.58% |  |  |
| **Category 1** |  |  |  |  |
| Under 50 | 14 | 29.17% |  |  |
| Over 50 | 34 | 70.83% |  |  |
| **Category 2** |  |  |  |  |
| Male, Under 50 | 5 | 10.42% |  |  |
| Male, Over 50 | 12 | 25.00% |  |  |
| Female, Under 50 | 9 | 18.75% |  |  |
| Female, Over 50 | 22 | 45.83% |  |  |
| Total | 48 |  |  |  |

## DISCUSSION

In this study, we present a semi-supervised Random Forest model for wrist and hip worn monitors to classify free-living physical activity intensity. The two-stage classifier that first classified activity type and then activity intensity showed comparable estimates between the two placements and overall time in each physical activity category that did not differ from ground-truth time.

The machine learning classifiers re-trained on free-living data accurately estimated light, moderate and vigorous intensity between both device placements. These findings demonstrate that similar estimates of physical activity intensity can be correctly classified for wrist and thigh placements when using semi-supervised techniques. The new intensity classification may have wider applications to combine datasets with wearables worn on different placements for utility in future research.

In particular, classification of physical activity intensity that is robust to different wear placements will allow for to design and evaluate studies examining the health effects of moderate-vigorous physical activity or sitting time regardless of if the device is worn on the wrist or thigh. Notably, the wrist classifier trained on the dominant wrist showed consistent performance when tested on the non-dominant wrist, when participants wore both simultaneously during the free-living sessions. Future research should explore the comparisons of activity type classifications between different wear placements, particularly as there are significant biomechanical differences and a wider range of movement degrees for the wrist compared to the thigh that may have a different impact and subsequent signal processing techniques that need to be used for activity type classification than activity intensity classification.

## Data Availability

All data produced in the present study are available upon reasonable request to the authors

## References

1. Ellis, K., Kerr, J., Godbole, S., Staudenmayer, J., & Lanckriet, G. (2016). Hip and wrist accelerometer algorithms for free-living behavior classification. Medicine and science in sports and exercise, 48(5), 933.

2. Herrmann SD, Willis EA, Ainsworth BE, et al. 2024 Adult Compendium of Physical Activities: A third update of the energy costs of human activities. Journal of Sport and Health Science. 2024;13(1):6–12. doi:10.1016/j.jshs.2023.10.010

3. McKenzie TL. Use of direct observation to assess physical activity. Physical activity assessments for health-related research. 2002;179:195.

4. Kerr, J., Patterson, R. E., Ellis, K., Godbole, S., Johnson, E., Lanckriet, G., & Staudenmayer, J. (2016). Objective assessment of physical activity: classifiers for public health. Medicine and science in sports and exercise, 48(5), 951.

5. Ahmadi M, O’Neil M, Fragala-Pinkham M, Lennon N, Trost S. Machine learning algorithms for activity recognition in ambulant children and adolescents with cerebral palsy. Journal of NeuroEngineering and Rehabilitation. 2018/11/15 2018;15(1):105. doi:10.1186/s12984-018-0456-x

6. Korshøj M, Skotte JH, Christiansen CS, et al. Validity of the Acti4 software using ActiGraph GT3X+accelerometer for recording of arm and upper body inclination in simulated work tasks. Ergonomics. 2014;57(2):247–53. doi:10.1080/00140139.2013.869358

7. Skotte J, Korshøj M, Kristiansen J, Hanisch C, Holtermann A. Detection of physical activity types using triaxial accelerometers. Journal of physical activity and health. 2014;11(1):76–84.

8. Ray, E. L., Sasaki, J. E., Freedson, P. S., & Staudenmayer, J. (2018). Physical activity classification with dynamic discriminative methods. Biometrics, 74(4), 1502–1511.

9. Breiman L. Random Forests. Machine Learning. 2001/10/01 2001;45(1):5–32. doi:10.1023/A:1010933404324

10. Prasetiyowati MI, Maulidevi NU, Surendro K. Determining threshold value on information gain feature selection to increase speed and prediction accuracy of random forest. Journal of Big Data. 2021/06/05 2021;8(1):84. doi:10.1186/s40537-021-00472-4

11. Stikic M, Van Laerhoven K, Schiele B. Exploring semi-supervised and active learning for activity recognition. IEEE; 2008:81–88.

